# Scaling analysis of COVID-19 spreading based on Belgian hospitalization data

**DOI:** 10.1101/2020.03.29.20046730

**Authors:** Bart Smeets, Rodrigo Watté, Herman Ramon

## Abstract

We analyze the temporal evolution of accumulated hospitalization cases due to COVID-19 in Belgium. The increase of hospitalization cases is consistent with an initial exponential phase, and a subsequent power law growth. For the latter, we estimate a power law exponent of ≈ 2.2, which is consistent with growth kinetics of COVID-19 in China and indicative of the underlying small world network structure of the epidemic. Finally, we fit an SIR-X model to the experimental data and estimate the effect of containment policies in comparison to their effect in China. This model suggests that the base reproduction rate has been significantly reduced, but that the number of susceptible individuals that is isolated from infection is very small. Based on the SIR-X model fit, we analyze the COVID-19 mortality and the number of patients requiring ICU treatment over time.

## Introduction

As of March 2020, the epidemic of new corona virus disease (COVID-19) is rapidly spreading throughout European countries. Some countries, such as Italy and Spain, have witnessed an explosion in cases, quickly saturating the treatment capacity of hospitals. To help steer governmental policies for containing the epidemic and to aid in the preparation planning of health services, understanding of the spreading behavior of COVID-19 through the population is critical.

Studies on the outbreak of COVID-19 in the Hubei province and the rest of mainland China show that the temporal evolution of confirmed cases can be classified in three distinct regimes: 1) an initial exponential growth phase, 2) an extended phase of power law growth kinetics indicative of a small world network structure, with a universal growth exponent of µ ≈ 2.1, and 3) a slow inflection to a plateau phase, following a parabolic profile in double logarithmic scale [1]. The roughly quadratic growth can be explained by considering the population as a two-dimensional planar network where the infected population only grows in the periphery of isolated ‘patches’ of infection [2]. The observed final inflection is not to be confused with the saturation of a logistic growth curve, which arises due to negative feedback as the number of susceptible people decreases with spreading of the infection. This effect is unlikely to contribute in the Chinese case, since even pessimistic estimates of the total number of COVID-19 cases stay very small compared to the total population. More likely, this effect can be attributed to extreme containment measures enacted in China. These measures disconnect the social network structure, producing caging effects that sufficiently slow down the spreading below a reproduction number of 1.

A popular epidemiological model is the SIR model, which is based on the formulation of ordinary differential equations for the number of susceptible (S), Infectious (I) and Recovered, or Removed (R) individuals [3]. This model was recently extended to include symptomatic quarantined individuals (X), resulting in the ‘SIR-X’ model, which was successfully applied to predict the spreading kinetics and assess containment policies for COVID-19 in China [4], and is currently being used to monitor the number of confirmed COVID-19 cases in various countries [5].

In Belgium, policies to contain the spreading of COVID-19 have proceeded in multiple phases. Initially, in phase I, strong quarantine measures were imposed on detected and suspected individuals who traveled from at-risk regions. In case of confirmed infections, their recent history of contacts was traced back and these individuals were quarantined as well as tested for COVID-19. Phase II included fast testing and quarantine of all individuals that exhibit symptoms. In phase III, drastic societal containment strategies are enacted. Regardless of symptoms, individuals are to minimize any social interactions. Due to restricted testing capacity, tests are only performed on individuals that exhibit severe symptoms. An important consequence of this strategy is that the number of confirmed cases can be heavily biased by shifting testing capacity and testing priorities. As an alternative, we propose that the accumulated number of hospitalized individuals is a good indicator for the number of actual COVID-19, albeit with a shift in time. This temporal shift is roughly equal to the combination of the mean incubation time (*≈* 5 days) and the average time from the onset of symptoms to hospitalization (*≈* 2 days) [6].

## Methods

### Data

Data is obtained from publicly available numbers on current hospitalization (*H*), current number of ICU patients (*ICU*), accumulated number of deaths (*D*) and number of individuals released from the hospital (*R*). These statistics are made public on a daily basis starting from March 13th 2020, based on data from more than 99% of Belgian hospitals [7]. For each day, the accumulated number of hospitalizations was computed as *H*_*a*_ = *H* + *R* + *D*. Here, we include data up to the 28th of March (release on the 29th). Throughout the analysis, dates shown indicate the date of publication of new data, and the ‘day’ scale counts the number of days starting from March 12th.

### Model

We use the SIR-X model introduced by Maier and Brockmann (2020) to simulate the hospitalization data. This model is based on the following ODEs [4]:

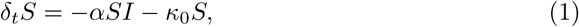

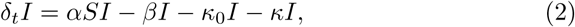

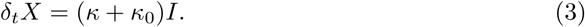

Here, *α* is the infection rate, *β* the recovery rate, *κ* the removal rate of symptomatic infected individuals, and *κ*_0_ the containment rate of both *S* and *I* populations. Originally, it is assumed that the fraction *X* is proportional to the number of confirmed infected cases. We will assume that *X* is proportional to the number of hospitalized cases, estimating that around 5% of infected (self-isolating) cases will be hospitalized, and that this occurs with a time delay of 2 days (average time between onset of symptoms and hospitalization). The precise proportionality of this scaling does not affect the further outcome of our analysis. The SIR-X model measures the effectiveness of isolation strategies through the ‘leverage factor’

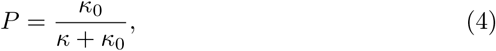

and the ‘quarantine probability’

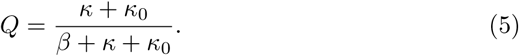

*P* is a measure for how strong isolation policies affect the general public in comparison to quarantine measures on infected individuals. *Q* is the probability that an infected individual is (self)quarantined. Moreover, it allows for the formulation of an effective reproduction number *R*_0,eff_ = *α/*(*β* + *κ* + *κ*_0_), which is always smaller than the basic reproduction number in free unconstrained growth *R*_0,free_ = *α/β*. Parameters *α* and *β* represent intrinsic properties of infectiousness and are not varied, but fixed at *α* = 0.775 and *β* = 0.125, corresponding to a recovery time of 8 days, and a free reproduction number of *R*_0,free_ = 6.2, as was assumed by the original authors [4]. The free parameters during the fitting procedure are *κ, κ*_0_ and *I*_0_*/X*_0_, the initial fraction of infectious individuals.

### Fitting routine

Fits are performed using the Levenberg-Marquardt least squares methods. During this procedure Eqs. (1-3) are integrated using the Dormand-Prince method, which uses a fourth-order Runge-Kutta method. The implementation of the fitting routine for the SIR-X model was kindly provided by the original authors [4]. The fitting of power law models was performed in double logarithmic space, and discarded the first five data points (March 12-16), which account for the exponential behavior.

## Results

Fig. 1 shows *H*_*a*_, *D* and *R* as a function of time. The accumulated hospitalization *H*_*a*_ showcases two distinct regimes: an initial exponential growth phase and a power law growth phase where *H*_*a*_ *∝ t*^*µ*^. For the latter, we estimate a fractal exponent *µ* = 2.22. The number of deaths *D* follows a power law growth 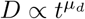 with *µ*_*d*_ = 3.27. As of March 29th, no significant deviations of neither *H*_*a*_ nor *D* from the power law growth can be observed. Furthermore, Fig. 1 shows the predicted accumulated hospitalization *X* and infectious population *I* from a fit using the SIR-X model. The parameters that fit the observed growth of *H*_*a*_ are listed in Table 1. The estimated value of *κ*_0_, the containment rate of the whole population is very close to zero. Consequently, the public containment leverage *P* is low as well. The quarantine probability is estimated at a value of *Q* = 0.780. Furthermore, there is a strong reduction of the reproduction number, with an effective reproduction number of *R*_0,eff_ = 1.36, much smaller than the unrestrained reproduction number *R*_0,free_ = 6.2. Finally, the SIR-X model predicts that the maximal number of infectious individuals occurs around April 12.

**Table 1:** Fit parameters of power law (*µ* and *µ*_*d*_), and SIR-X model (Eqs. 1-3).

| Parameter | Estimate | Std. Error |
| --- | --- | --- |
| $\mu$ | 2.216 | 0.040 |
| $\mu_d$ | 3.272 | 0.090 |
| $\kappa_0$ | 0.006 | 0.005 |
| $\kappa$ | 0.438 | 0.040 |
| $P$ | 0.014 | 0.011 |
| $Q$ | 0.780 | 0.015 |
| $R_{0,\text{eff}}$ | 1.362 | 0.096 |

**Figure 1:**
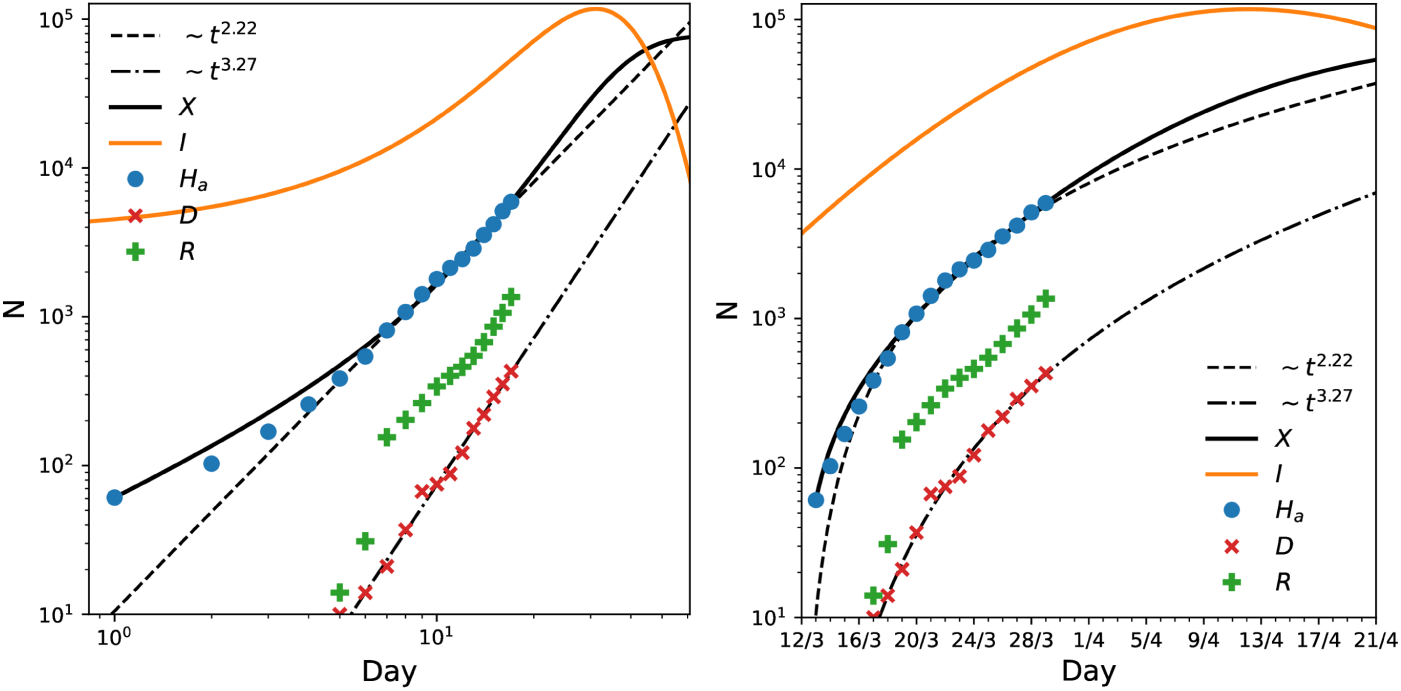
Temporal evolution of accumulated number of hospitalized individuals *H*_*a*_, including number of deaths *D* and accumulated number of hospital releases *R*. The same data is shown on the right on a linear time scale. Dashed lines indicate power law fits for *H*_*a*_ and *D*, and *X* and *I* indicate the predicted accumulated hospitalized cases and number of infected individuals from fitting the SIR-X model.

Fig. 2(a) shows the number of accumulated hospitalizations as well as deaths due to COVID-19 in comparison to the fitted SIR-x model. Setting an average mortality of 15% for all hospitalized cases [9], we find that the SIR-X model coincides with the number of deaths when including a temporal delay of only*≈* 5 days. Assuming an average hospitalization time of 12 days, and that between 15% and 20% of currently hospitalized patients require intensive care treatment (ICU), we predict based on the SIR-X model the temporal evolution of the current number of patients in ICU – Fig. 2(b-c). These assumptions align with the observed current number of ICU patients. For the estimated SIR-X model parameters, the number of ICU patients will peak around April 20th. The peak count of ICU patients greatly varies with the average ICU retention time, but will peak at significantly higher values than the current ICU capacity in Belgian hospitals of 2650 beds, Fig. 2(d).

**Figure 2:**
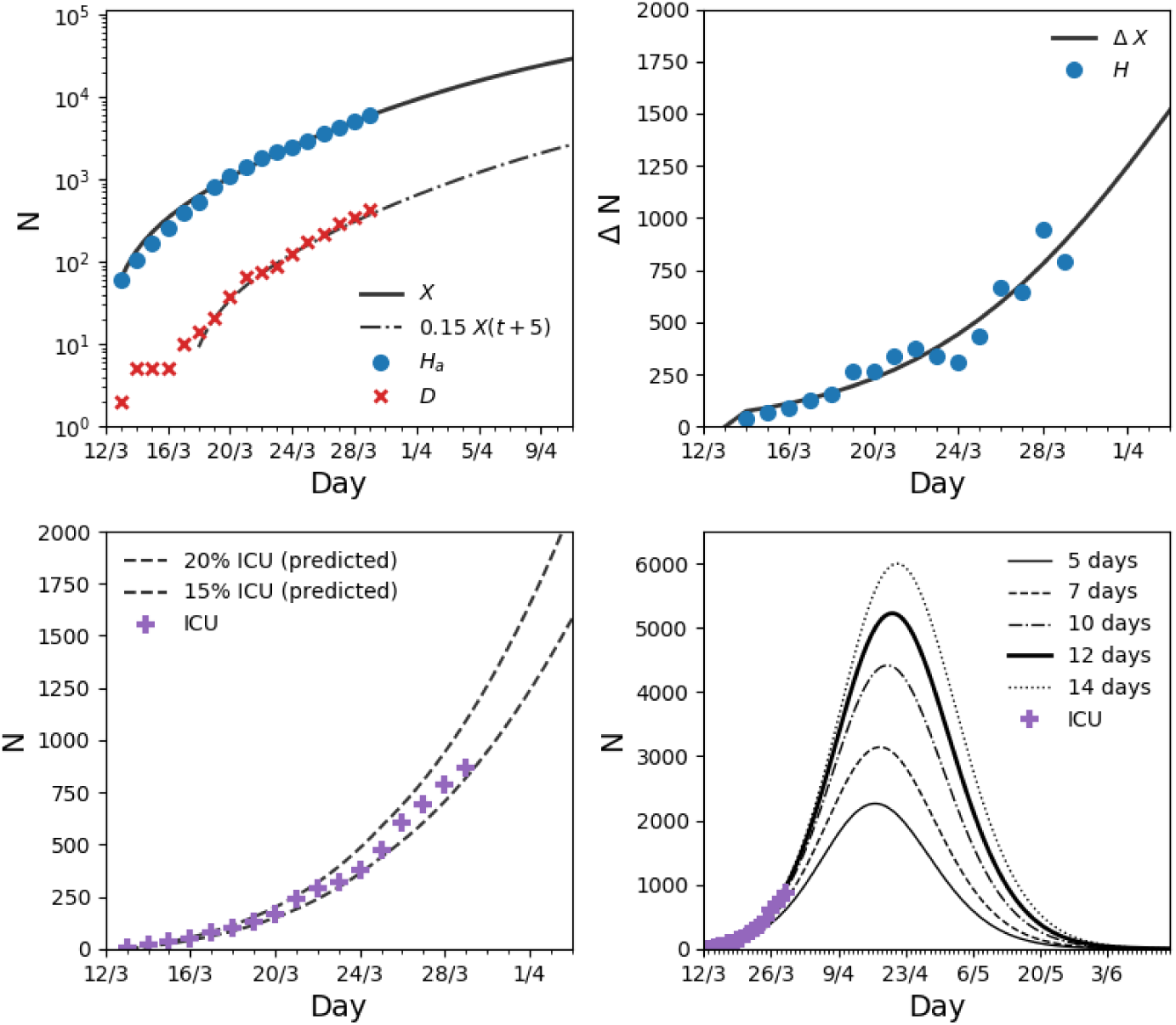
Analysis based on SIR-X model fit. **(a)** Predicted number of accumulated hospitalizations from SIR-X model, together with observed (*D*) and predicted mortality, where we have assumed a time delay of 5 days from hospitalization to death, and a mortality of hospitalized individuals of 15%. **(b)** Daily difference (Δ*N*) in current hospitalization (*H*) and predicted hospitalization computed as *X*(*t*) *− X*(*t−*12), assuming a average hospitalization time of 12 days. **(c)** Current number of patients treated in intensive care units (ICU), compared to predictions from the SIR-X model, assuming that 15% or 20% of hospitalized patients require intensive care. **(d)** Longer-time extrapolation assuming 17.5% ICU shows a peak of ICU patients around April 20th. Different curves show varying ICU retention times. The shorter the ICU retention time, the lower and earlier the ICU peak will be.

## Discussion

The time evolution of the accumulated number of hospitalizations in Belgium follows similar power law scaling (*µ* = 2.22) as the growth of disease in different regions of China, with papers reporting exponents of 2.1 [4], 2.25 [1] and 2.5 [8]. This approximately quadratic growth indicates a small-world network structure with mostly local interactions through which the spreading of the infection occurs. This is consistent with the observation based on data from Belgian telecom operators, which show that more than 80% of Belgians have stayed within their own commune (postal code) for the last two weeks, and that individual displacements of over 40 km have been reduced by 90%. At the time of this writing, no significant deviation from this power law scaling has been observed for the accumulated Belgian hospitalization data. This indicates that the current social network is still sufficiently well connected to continue local spreading of the disease. In part, these local network links could be attributed to the infection of direct family members. Without extensive testing, many infected people will be locked into their homes and pose a contamination risk for their families. In a severe lock-down, this effect should be controlled within a few days. Hence, other factors may contribute to the continuation of the power law scaling.

Another spreading mechanisms could be local supermarkets, where in spite of extensive safety measures, significant spreading of the highly infectious corona virus could occur. A solution can be to enforce a more rigid approach, in which supermarkets are viewed as local distribution centres. Most supermarkets already provide online shopping services, where people fill in their online shopping carts. This would allow for an optimal spreading of customers, and/or would make it possible to deliver groceries in a drive-through system. The result would be a large reduction of the small-world connectivity, resulting in a lowering of the exponents, thereby further flattening the curve.

Since the current scaling behavior of *H*_*a*_ still closely follows the algebraic growth regime, it is very difficult to make accurate predictions on when the inflection point away from this regime will occur. As a rule-of-thumb, reliable prediction capacity does not extend a period of about 3 days. The SIR-X model predicts that *H*_*a*_ will start to plateau around 40 days after the initial day. The parameters from the SIR-X model fit suggest a very low value for the containment rate of both infected and susceptible individuals *κ*_0_. In other words, containment measures have only a weak effect on removing healthy individuals from the susceptible population. On the other hand, the removal rate of symptomatic individuals is much higher, leading to a strongly decreased effective reproduction number, and a moderately high quarantine probability. These SIR-X model parameters are somewhat similar to values estimated for the Beijing region of China [4]. When extrapolating the number of deaths using the SIR-X model, the predicted death toll due to COVID-19 will exceed 10^3^ by April 3rd. The SIR-X model for accumulated hospitalizations is compatible with the current number of patients in intensive care when assuming that between 15% and 20% of hospitalized patients need ICU treatment, and that the average retention time in ICU is around 12 days. Extrapolation with these parameters predicts a peak in number of ICU patients around April 15th, with the number of ICU patients exceeding the capacity of the Belgian healthcare system of 2650 beds.

Based on Fig. 2(a,c-d), one can conclude that the model matches very well with the total number of hospitalisations over time. This experimentally determined parameter is a numerical integration of the number of new cases each day. This integration has the important advantage of averaging out the noisiness in the day-to-day reporting – Figure 2(b). Belgian media have reported that some hospitals have published their numbers with a time-delay of one day, which has a large impact on the visualisation of the results in a linear scale.

It should however be emphasized that these predictions are highly sensitive to the estimated parameters of the SIR-X model [4]. Furthermore, the model assumes that these constants will not further change in time. In reality, the effect of containment and isolation measures may occur gradually and with a significant time delay.

## Data Availability

All data is publicly available and can be obtained from the Belgian platform for infectious diseases (epidemio). All analysis code is available on github.

https://github.com/smeetsbart/covid-hospitalization-belgium

https://github.com/benmaier/COVID19CaseNumberModel

